# Advertising ultra-processed foods around urban and rural schools in Kenya

**DOI:** 10.1101/2024.09.10.24313437

**Authors:** Caroline H. Karugu, Gershim Asiki, Milka Wanjohi, Veronica Ojiambo, Richard Sanya, Amos Laar, Michelle Holdsworth, Stefanie Vandevijvere, Charles Agyemang

**Affiliations:** Chronic Diseases Management Unit, African Population Health Research Center, Nairobi, Kenya; Department of Public and Occupational Health, Amsterdam Public Health, University of Amsterdam Medical Centers, Amsterdam, the Netherlands; Department of Women’s and Children’s Health, Karolinska Institute, Stockholm, Sweden; Department of Public & Global Health, University of Nairobi, Kenya; School of Public Health, University of Ghana, Accra, Ghana; UMR MoISA (Montpellier Interdisciplinary Centre On Sustainable Agri-Food Systems), (Univ Montpellier, CIRAD, CIHEAM-IAMM, INRAE, Institut Agro, IRD), Montpellier, France; Sciensano, Service of Lifestyle and Chronic Diseases, Brussels, Belgium; Division of Endocrinology, Diabetes, and Metabolism, Department of Medicine, Johns Hopkins University School of Medicine, Baltimore, MD, USA

**Keywords:** Food marketing, Outdoor advertisements, Healthy foods, Unhealthy’ foods, Ultra-processed foods, Sugar-sweetened beverages, School neighborhoods, Food environments

## Abstract

**Background:** Marketing of ultra-processed foods (UPFs) can influence purchase intentions and consumption of such foods, especially among children. There is limited evidence on the extent to which UPFs are marketed around schools in low-and middle-income countries (LMICs), including Kenya. We assessed the extent, content, and type of advertising of ultra-processed/unhealthy foods around schools in urban and rural settings in Kenya.

**Methods:** This cross-sectional study assessed advertising of food and beverages within a 250m radius around schools in Kenya between June to July 2021. We conducted the study in three counties (Nairobi-urban, Mombasa-coastal urban city, and Baringo-rural). Each county was stratified into high and low socioeconomic status (SES) sub-counties. Within each, we randomly selected schools and collected detailed information on advertising around the schools. The information gathered included the location, type of food and beverage product advertised, and the promotional techniques used. We used the NOVA classification and International Network for Food and Obesity/NCDs Research Monitoring and Action Support (INFORMAS) methods to categorize the food and beverages advertised based on processing level and core (healthy)/non-core(unhealthy) groups. We determined the extent of advertisements using descriptive analysis frequencies, and median (interquartile ranges). Poisson regression was used to determine the factors associated with UPF advertisements.

**Results:** In total, 2300 food and beverage advertisements were mapped around the 500 schools. There was a higher median number of advertisements in urban areas (median=25, Interquartile range (IQR)=25,160) compared to rural areas (median=10, IQR= 4, 13). Of these advertisements, approximately 48.0% were UPFs. The most common promotional strategy used was cartoon and company-owned characters, while the most frequent premium offer was price discounts. In the multivariate analysis, there was a significantly higher rate of advertisements of UPFs in Baringo County (prevalence rate ratios (PRR): 1.17, 95% CI: 1.01-1.36) compared to the urban Nairobi County, and in lower compared to higher SES settings (PRR: 1.10, 95% CI: 1.01-1.20).

**Conclusion:** UPFs are frequently advertised around schools with promotional strategies that may be appealing to children. There is a need to restrict the marketing of unhealthy foods around schools to children in Kenya.

## Introduction

Overweight and obesity are a growing global challenge largely driven by obesogenic food environments and the consumption of unhealthy foods, as well as insufficient physical activity (1). Obesity prevalence is projected to increase over time, particularly in low and middle-income countries (LMICs) (2). A major challenge in LMICs is the double burden of malnutrition in children. Overweight and obesity and undernutrition in childhood are all associated with an increased future risk of non-communicable diseases in adulthood (3).

The economic and industrial transition in various settings has impacted the food environments that children and adults are exposed to substantially. There is an increasing trend in the marketing of unhealthy foods such as ready-to-eat snacks and sweetened drinks and beverages targeting children and adolescents (4–6). Children are progressively exposed to unhealthy foods and beverages by industries targeting settings and platforms such as schools, the traditional media including TV and radio, and, social media (3,7,8). Several persuasive techniques such as cartoons, company-owned characters, celebrity brand endorsements, gifts and collectibles, price discounts, and game application downloads are used to entice children toward the purchase and consumption of unhealthy foods(9,10). Marketing of unhealthy foods influences dietary habits, attitudes, and preferences toward unhealthy foods, and increased consumption of the marketed food and beverages (6). In both HICs and LMICs, there is an increasing trend in advertising unhealthy foods and beverages targeting children (11).

Various studies have been conducted in both LMICs and HICs illustrating the extent of marketing health-harming food products to children and adolescents (12–18). Studies conducted in New Zealand and Australia representing different socioeconomic deprivation areas showed that the majority of the advertised food and beverage products were prohibited by the World Health Organization (WHO) nutrition profile model and were mostly soft drinks and alcoholic drinks(12,13). In the Philippines and Guatemala, studies assessing outdoor advertisements around school neighborhoods showed a high density of advertisements around schools and a high level of advertisements of unhealthy foods that target children (19,20). A study conducted in Uganda’s capital-Kampala-around school neighborhoods showed that more than 80% of advertisements were for sugar-sweetened beverages, especially in urban settings(21). In Ghana, approximately 69% of the food and beverage products advertised were in the ultra-processed foods (UPFs) category, with the cartoon and company-owned characters as the main promotional characters used (18). Most of the studies on marketing around school environments have focused on urban neighborhoods (18,21).

In this study, we assessed the extent, content, type, and power of advertising of ultra-processed/unhealthy foods around schools in both urban and rural settings in Kenya. We further explored other factors associated with the marketing of UPFs.

## Materials and methods

### Study design

This was a cross-sectional study conducted in Kenya from June 2021 to July 2021, which assessed the food and beverage marketing around primary and secondary schools. The study adopted the International Network for Food and Obesity/NCDs Research Monitoring and Action Support (INFORMAS) methodology, the outdoor food advertisement protocol in particular(22).

### Study setting

Three counties (Nairobi, Mombasa, and Baringo) were selected to represent different contexts including urban and rural, low and high socio-economic status (SES) within urban settings, and a tourist destination in Kenya (Figure 1). Each county was stratified into low and high SES sub-counties using poverty-level data from the 2019 Kenya National Bureau of Statistics (KNBS) estimates (23). In Nairobi County Westlands (higher SES), Langata (higher SES), Embakasi (lower SES), Kibra (lower SES), and Mathare (lower SES) were selected. In Mombasa county, Mvita (Higher SES), and Kisauni (lower SES) sub-counties were selected. Lastly, in Baringo county, we included Mogotio (higher SES), and Baringo North (lower SES) sub-counties.

**Figure 1:**
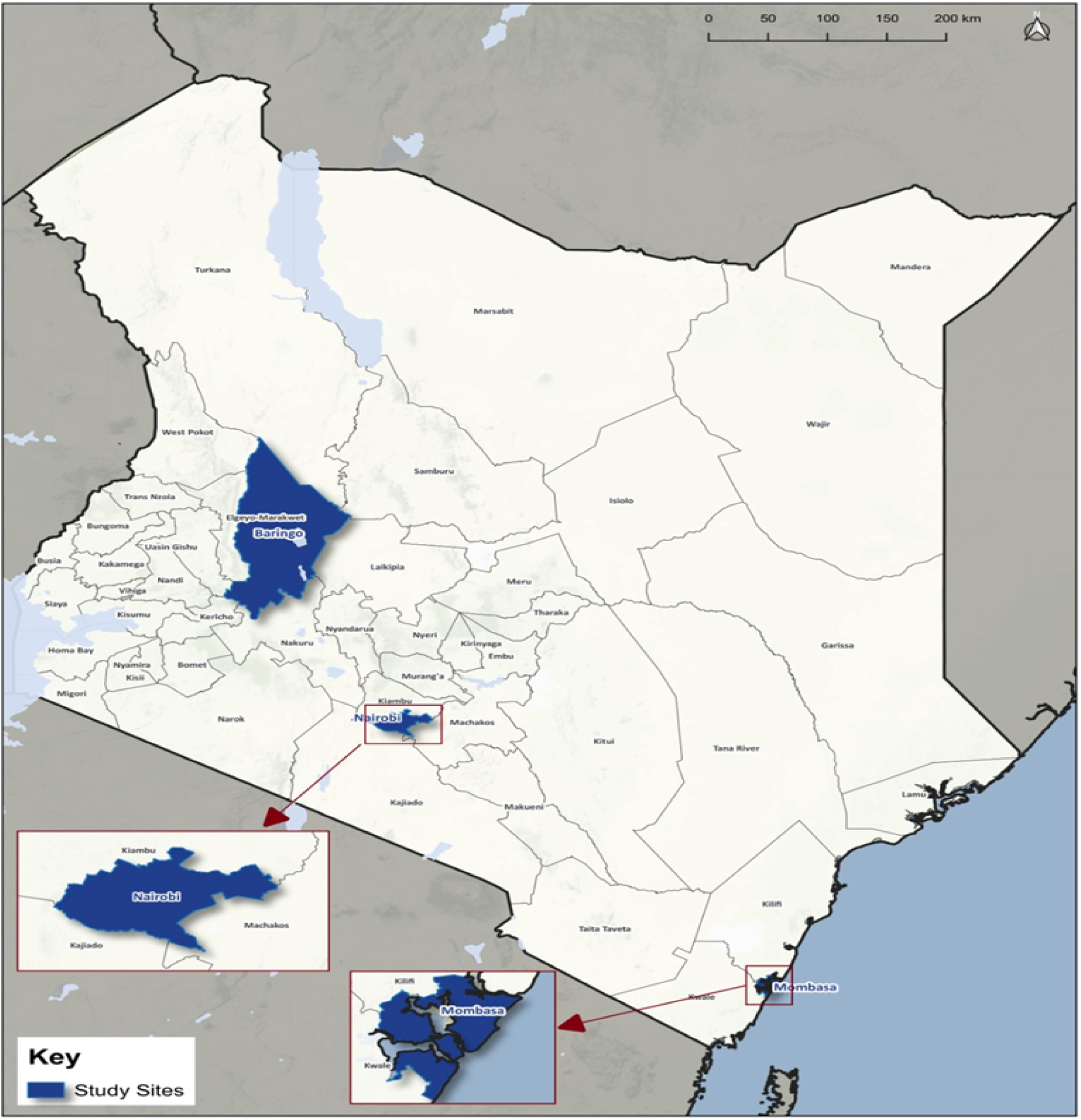
Map of Kenya showing the counties selected for assessing food availability and marketing at modern food retailers in 2021.

### Sample size and sampling

We employed a multistage stratified sampling technique in this study. First, we estimated a sample size of 500, considering a prevalence of 23% of unhealthy food advertisements in deprived-resource settings (24), a power of 80%, and a margin of error of 0.05. We further adjusted this sample size considering a 10% nonresponse rate in all the settings. The 500 schools were further randomly allocated proportionately to the number of schools by level (primary or secondary and type (private or public) in each county and sub-county using the stratified sampling method. The list of schools by location, level, and type was provided by the Ministry of Education.

### Data collection procedures

After obtaining ethical approvals from the AMREF-Health Ethics and Scientific Review Committee in Kenya (approval number: ESRC/P901/2020), and permission from the school heads, the research team comprising two data collectors per school at a time was deployed to the selected schools. They were trained to identify the required distance/radius from the main school gates and entrances and collect data on the advertisements within the defined radius (24), as per the INFORMAS outdoor advertisements protocol used in previous studies in LMIC settings (18,22).

### Food marketing assessments/ Study variables

The details of the schools including the school name, school type (private, public, primary school, secondary school), school setting (urban, rural, peri-urban), school population, and number of school entrances were recorded. All the advertisements within the pre-determined Euclidean radius around the schools were recorded. This involved first identifying the location of the food and beverage advertisement; whether the product was placed in food shops, buildings, on the roadside, bus shelter/bus stage, or mobile karts. The format of advertisements (billboards, posters/banners, standing signs, painted walls and buildings, digital signs, and store merchandising) n was also recorded. The size of the advertisements was determined either as small (> A4 but <1.3m X 1.9m), medium (<1.3m X 1.9m but < 2.0m X 2.5m), or large (2m X 2.5m). Further, we assessed the content of the advertisement (logos/company pictorials, text, combined logos, and text) and the number of food products in the individual advertisement (single-food product type, two-food product type, three-food product type, and >3 food product type). The individual food item advertisement was recorded and the corresponding promotional characters and premium offers were used. The promotional characters include cartoon or company-owned characters, famous sports persons, movie titles, or images with kids. The premium offers included price discounts, limited stock editions, and buy one get one free.

### Food classification

The food and beverage products advertised around school neighborhoods were categorized based on the INFORMAS food categories(25), and the NOVA classification systems(26). Alcoholic beverages were not included in the food categorizations and were reported as a separate group. The INFORMAS food categorization classifies foods into core (healthy) and non-core (unhealthy) food categories (25), (26), (27). Examples of core foods are fruits and vegetables, while non-core foods are unhealthier food options such as sugar-sweetened beverages. The NOVA food classification system classifies foods into i) unprocessed/minimally processed which are foods in their raw state that have undergone no industrialization processes and have no added sugars and salts ii) processed culinary ingredients, iii) processed foods, and iv) Ultra-processed foods(UPFs)(28). Additionally, the food categories were further subcategorized into a binary variable: UPFs and not UPFs.

### Statistical analysis

The analysis was guided by the INFORMAS protocol (28). Descriptive statistics were used to show the distribution of foods advertised by counties, SES areas, and other characteristics such as school types and levels and level of urbanization. We used median and interquartile ranges to assess the extent of advertisements around schools. The outcome of the study was the extent of advertisements of core (healthy) and non-core (unhealthy) foods, and UPFs advertisements around schools. The predictor variables included counties, SES levels, rural/urban settings, school type, school population, and the gender of the student population within the schools. The multivariable Poisson regression models were used to determine the association between the count of food and beverages advertised around the schools and other characteristics such as SES status and school type. We adjusted for clustering within the individual sub-counties in the Poisson regression models to account for the differences in the settings of these regions. Post-estimations were conducted to assess the validity of the models.

### Ethical approvals

The study protocol was approved by the AMREF-Health Ethics and Scientific Review Committee in Kenya (ESRC/P901/2020) and the National Commission for Science, Technology, (NACOSTI-P/22/19104). Permission and mobilization were done to the school heads and subcounty level administration.

## Results

### School characteristics description

Advertising was assessed around all 500 schools: Nairobi county schools (n=215, 43.0%), Baringo county schools (n=173, 34.6%), and Mombasa county schools (n=112, 22.4%). More than 50% of the schools were public schools, while 78.4% were primary schools (**Table 1**). Nearly two-thirds of the schools were from urban settings while most schools (>90%) were mixed gender (both male and female pupils). According to the SES assessment of the schools, 60.0% of them were in the lower SES (**Table 1**). The Mean (SD) school population in all the counties was 384.2(429.1) pupils. The schools had entrances ranging from 1 to 8.

**Table 1:** School characteristics in Nairobi, Mombasa, and Baringo Counties.

| Variable | Description | Frequency<br>(Percentage) (N=500)<br>n (%) |
| --- | --- | --- |
| **County** |  |  |
|  | Nairobi | 215 (43.0) |
|  | Mombasa | 112 (22.4) |
|  | Baringo | 173 (34.6) |
| **School ownership** |  |  |
|  | Private school | 238 (47.6) |
|  | Public school | 262 (52.4) |
| **School type** |  |  |
|  | Primary school | 392 (78.4) |
|  | Secondary school | 108 (21.6) |
| **School setting** |  |  |
|  | Urban | 323 (64.6) |
|  | Rural | 177 (35.4) |
| **Gender of pupils in the schools** |  |  |
|  | Boys school | 20 (4.0) |
|  | Girls school | 15 (3.0) |
|  | Mixed school | 465 (93.0) |
| **School population** |  |  |
|  | Mean (SD), range | 384.2 (429.1), (114, 3600) |
| **School SES area** |  |  |
|  | Lower SES | 300 (60.0) |
|  | Higher SES | 200 (40.0) |
| **Number of school entrances** |  |  |
|  | Mean (SD), range | 1.3 (0.8), (1, 8) |

### Extent of advertisements around schools

We observed a total of 2300 food and beverage advertisements, most of them from Nairobi county (n=1256; 54.8%), followed by Mombasa county (n=878; 38.2%), and the fewest from Baringo county (n=166; 7.2%). The food and beverage products were more advertised in Mombasa (Median=25, Interquartile range (IQR)= 12, 78) and Nairobi counties (Median=25, IQR=6, 69), and lower in Baringo county (Median=8, IQR; 5, 14) (**Table 2**). We observed a higher number of food and beverage advertisements in urban areas (Median=30, range= 25, 160), in higher SES areas (Median=25, IQR=12, 69), and around private primary schools (Median=19, IQR: 15, 92) (**Table 2**). We found a lower median number of advertisements around private secondary schools and public secondary schools.

**Table 2:** Median number of food and beverage advertisements around a 250 m radius around schools by county, SES levels, and school type in Nairobi, Mombasa, and Baringo counties.

| Variable | Characteristics | Number of schools (N=500) | Median advertisements around schools | (25th, 75-percentiles) |
| --- | --- | --- | --- | --- |
| **County** |  |  |  |  |
|  | Nairobi county | 215 | 25 | (12,78) |
|  | Mombasa county | 112 | 25 | (7, 69) |
|  | Baringo county | 173 | 8 | (5, 15) |
| **School setting** |  |  |  |  |
|  | Urban | 323 | 30 | (25, 160) |
|  | Rural | 177 | 10 | (4, 13) |
| **SES level** |  |  |  |  |
|  | Lower SES | 300 | 22 | (11, 83) |
|  | Higher SES | 200 | 25 | (12, 69) |
| **School type** |  |  |  |  |
|  | Private primary | 190 | 19 | (15, 92) |
|  | Public primary | 202 | 16 | (8, 35) |
|  | Private Secondary | 48 | 10 | (4, 14) |
|  | Public secondary | 60 | 6 | (3, 15) |

### Features of food and beverage advertisements

Table 3 shows the characteristics of food and beverage advertisements. In all the counties, 85.5% of all food and beverage advertisements were within food stores. Approximately 5.8% and 5.4% of these advertisements were located on buildings, and along the roads, respectively. Advertisements were mainly displayed as posters and banners (60.6%), followed by store merchandising (25.4%), and painted buildings/walls (8.4%). Most food and beverage advertisements were promoted as single products (61.8%), and 16.0% promoted more than three food product types (**Table 3**). The most common content of the advertisements (86.7%) was combined logos, pictorials, and text. About 46.5% of the advertisements were small-sized, and 38.2% and 15.2% were medium-and large-sized, respectively.

**Table 3:** Techniques for food advertising in/around schools in Nairobi, Mombasa, and Baringo Counties.

|  | *Description* | *Frequency (%)* |  |
| --- | --- | --- | --- |
| **Setting of advertisement** | Food shop | 1,966 | (85.5%) |
|  | Road | 125 | (5.4%) |
|  | Building | 133 | (5.8%) |
|  | Bus shelter/ bus stage/bus stop/train | 2 | (0.1%) |
|  | Mobile cart/stall or vending machine | 17 | (0.7%) |
|  | Other specify <sup>a</sup> | 57 | (2.5%) |
| **Format of advertisement** | Billboard | 86 | (3.7%) |
|  | Poster or banner | 1,393 | (60.6%) |
|  | Standing sign | 30 | (1.3%) |
|  | Painted building/wall | 192 | (8.4%) |
|  | Digital signs / LED | 10 | (0.4%) |
|  | Store merchandising | 583 | (25.4%) |
|  | Others specify <sup>b</sup> | 6 | (0.3%) |
| **Number of product types in advertisement** | None (only company/brand mentioned) | 82 | (3.6%) |
|  | 1 food product type | 1,422 | (61.8%) |
|  | 2 food product types | 210 | (9.1%) |
|  | 3 food product types | 219 | (9.5%) |
|  | > 3 food product types | 367 | (16.0%) |
| **Content of the advertisement** | Logos/Pictorial | 111 | (4.8%) |
|  | Text | 195 | (8.5%) |
|  | Combined logos, pictorials, and text | 1994 | (86.7%) |
| **Size of advertisement** | Small ( > A4 but <1.3m X 1.9m) | 1070 | (46.5%) |
|  | Medium (<1.3m X 1.9m but < 2.0m X 2.5m) | 878 | (38.2%) |
|  | Large (2m X 2.5m) | 352 | (15.2%) |

Notes: <sup>a</sup> **Other settings include: advertisements on gates, bars, restaurants, and liquor shops.**
<sup>b</sup> **Other formats include: Branded moving cars and calendar adverts**

### Types of foods and beverages advertised using INFORMAS and NOVA food categories

Table 4 shows the INFORMAS food categorization of the advertisements. About 46.0% of all the food and beverage advertisements were unhealthy food options, 46.4% were healthy foods, and approximately 5% were in the miscellaneous category. Approximately 3% of all the advertisements promoted alcoholic drinks and beverages. In all the counties the most frequently advertised food category was sugar-sweetened drinks, which falls in the unhealthy food category, accounting for more than 33.1% of all the advertisements. The most commonly advertised healthy food category was meat and meat alternatives, which contributed to 12% of all the advertisements (**Table 4**).

**Table 4:**
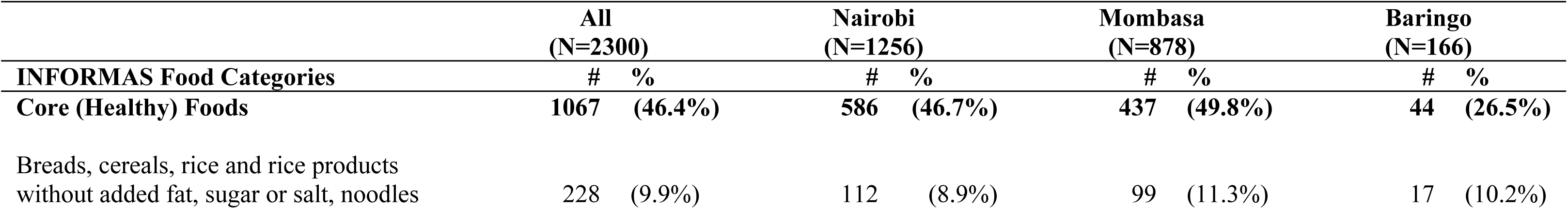
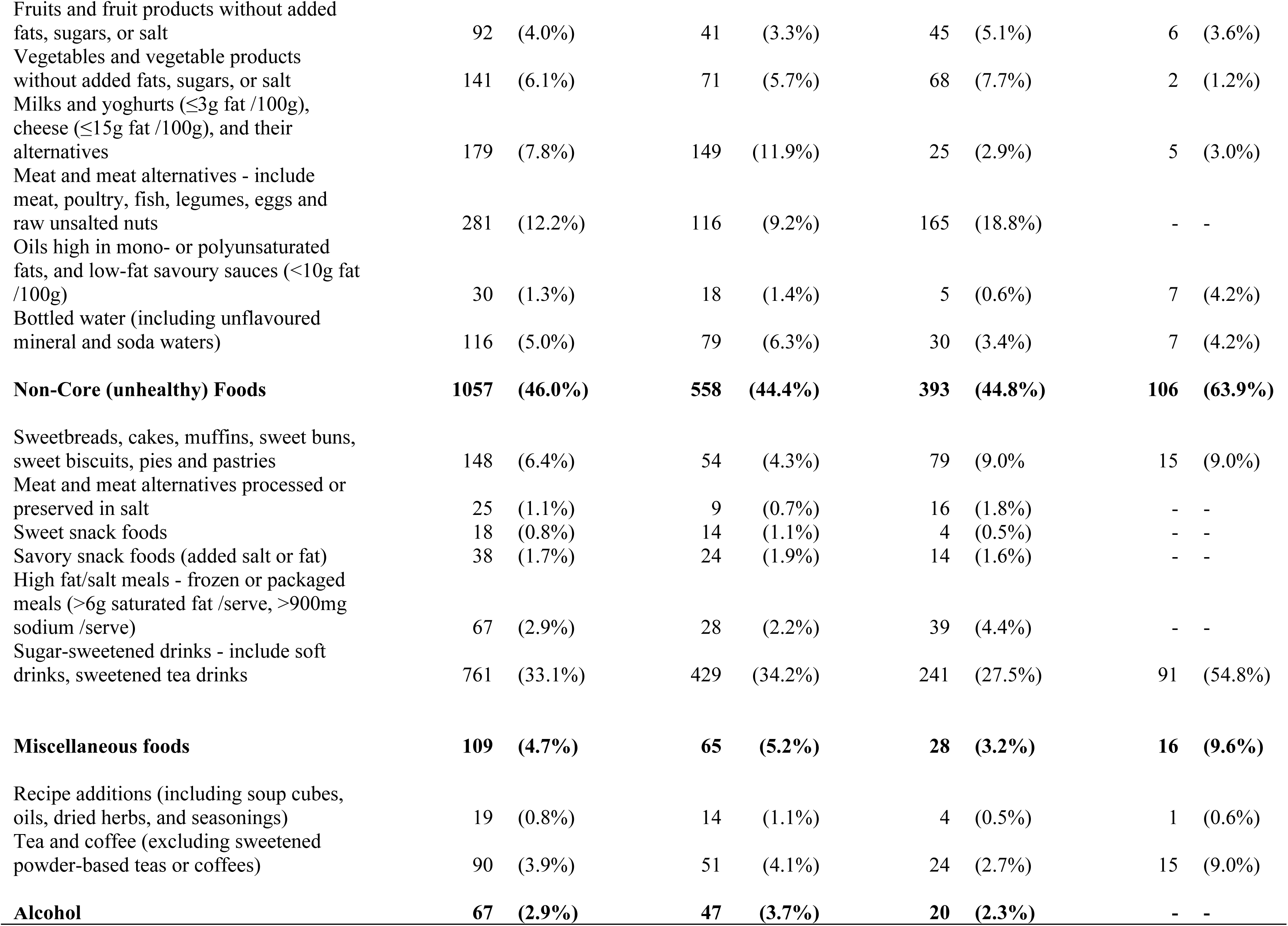
Food categories and distribution by the proportion of food advertisements in/ around schools in Nairobi, Mombasa, and Baringo Counties.

|  | All<br>(N=2300) |  | Nairobi<br>(N=1256) |  | Mombasa<br>(N=878) |  | Baringo<br>(N=166) |  |
| --- | --- | --- | --- | --- | --- | --- | --- | --- |
| INFORMAS Food Categories | # | % | # | % | # | % | # | % |
| Core (Healthy) Foods | 1067 | (46.4%) | 586 | (46.7%) | 437 | (49.8%) | 44 | (26.5%) |
| Breads, cereals, rice and rice products<br>without added fat, sugar or salt, noodles | 228 | (9.9%) | 112 | (8.9%) | 99 | (11.3%) | 17 | (10.2%) |

|  |  |  |  |  |
| --- | --- | --- | --- | --- |
| Fruits and fruit products without added fats, sugars, or salt | 92 (4.0%) | 41 (3.3%) | 45 (5.1%) | 6 (3.6%) |
| Vegetables and vegetable products without added fats, sugars, or salt | 141 (6.1%) | 71 (5.7%) | 68 (7.7%) | 2 (1.2%) |
| Milks and yoghurts (≤3g fat /100g), cheese (≤15g fat /100g), and their alternatives | 179 (7.8%) | 149 (11.9%) | 25 (2.9%) | 5 (3.0%) |
| Meat and meat alternatives - include meat, poultry, fish, legumes, eggs and raw unsalted nuts | 281 (12.2%) | 116 (9.2%) | 165 (18.8%) | - - |
| Oils high in mono- or polyunsaturated fats, and low-fat savoury sauces (<10g fat /100g) | 30 (1.3%) | 18 (1.4%) | 5 (0.6%) | 7 (4.2%) |
| Bottled water (including unflavoured mineral and soda waters) | 116 (5.0%) | 79 (6.3%) | 30 (3.4%) | 7 (4.2%) |
| **Non-Core (unhealthy) Foods** | **1057 (46.0%)** | **558 (44.4%)** | **393 (44.8%)** | **106 (63.9%)** |
| Sweetbreads, cakes, muffins, sweet buns, sweet biscuits, pies and pastries | 148 (6.4%) | 54 (4.3%) | 79 (9.0%) | 15 (9.0%) |
| Meat and meat alternatives processed or preserved in salt | 25 (1.1%) | 9 (0.7%) | 16 (1.8%) | - - |
| Sweet snack foods | 18 (0.8%) | 14 (1.1%) | 4 (0.5%) | - - |
| Savory snack foods (added salt or fat) | 38 (1.7%) | 24 (1.9%) | 14 (1.6%) | - - |
| High fat/salt meals - frozen or packaged meals (>6g saturated fat /serve, >900mg sodium /serve) | 67 (2.9%) | 28 (2.2%) | 39 (4.4%) | - - |
| Sugar-sweetened drinks - include soft drinks, sweetened tea drinks | 761 (33.1%) | 429 (34.2%) | 241 (27.5%) | 91 (54.8%) |
| **Miscellaneous foods** | **109 (4.7%)** | **65 (5.2%)** | **28 (3.2%)** | **16 (9.6%)** |
| Recipe additions (including soup cubes, oils, dried herbs, and seasonings) | 19 (0.8%) | 14 (1.1%) | 4 (0.5%) | 1 (0.6%) |
| Tea and coffee (excluding sweetened powder-based teas or coffees) | 90 (3.9%) | 51 (4.1%) | 24 (2.7%) | 15 (9.0%) |
| **Alcohol** | **67 (2.9%)** | **47 (3.7%)** | **20 (2.3%)** | **- -** |

**Table 5:** Healthiness of food and beverage categories advertised by school characteristics.

|  |  | *Core (healthy) foods* | *Non-core (unhealthy) foods* | *Miscellaneous* |  |
| --- | --- | --- | --- | --- | --- |
| *Variable* | *Characteristics* | *Median (25th, 75 percentile)* | *Median (25th, 75 percentile)* | *Median (25th, 75 percentile)* | *P-value* |
| **County** |  |  |  |  | 0.000 |
|  | Nairobi county | 79(56, 114) | 37.5(16.5, 47.5) | 32.5(23.25, 41.75) |  |
|  | Mombasa county | 45(26.25, 68.0) | 20(14.5, 39.0) | 14(9.0, 19.0) |  |
|  | Baringo county | 7(5.25, 7.0) | 15(53, 72) | 8(4.5, 11.5) |  |
| **School setting** |  |  |  |  | 0.000 |
|  | Urban | 136(99, 191) | 67(31, 101.5) | 46(31.5, 60.5) |  |
|  | Rural | 5(4, 5) | 12(6.5, 49.5) | 8.5(5.25, 11.75) |  |
| **SES level** |  |  |  |  | 0.000 |
|  | Lower SES | 73(37.5, 113.5) | 14(13.5, 52.5) | 29(20, 38) |  |
|  | Higher SES | 62(45, 103.5) | 26(18, 68.5) | 25.5(16.75, 34.25) |  |
| **School type** |  |  |  |  | 0.000 |
|  | Private primary | 95(63, 127.5) | 35(22.5, 56.0) | 25.5(17.75, 33.25) |  |
|  | Public primary | 25(17, 30.5) | 10(5, 35) | 17.5(11.25, 23.75) |  |
|  | Private secondary | 12(8.5, 15.5) | 11(4.75, 14.25) | 5.5(3.25, 7.75) |  |
|  | Public secondary | 11(8, 25.5) | 5(3, 7) | 6(4.5, 7.5) |  |

*Notes: P-Value from chi-square tests for checking differences in proportions of food categories by different variables such as SES levels*

In Nairobi county, 44.4% of the advertisements were noncore (median=37.5, IQR=16.5, 47.5) and 46.7% (median=79, IQR=56, 114) were core foods (**Table 4**, **5**). Conversely, 49.8% of the food and beverage advertisements in Mombasa county were healthy foods, while the unhealthy foods closely contributed to 44.8% of the food and beverage advertisements. In Baringo, unhealthy foods contributed approximately 63.9% (median=15, IQR=53, 72) of the food and beverage advertisements in the county (**Table 4**, **5)**. The sugar-sweetened beverages food category was the most commonly advertised in all three counties, contributing to approximately 34.2%, 27.5%, and 54.8% of the advertisements in Nairobi, Mombasa, and Baringo counties, respectively (**Table 4**). According to the bivariate analysis, there were significant differences in the marketing of core and non-core foods by counties, SES settings, school settings (urban/rural), and school type.

According to the NOVA food categorization, nearly equal proportions of the advertisements were for UPFs versus unprocessed/minimally processed (49% versus 48%) in all the counties. (**Figure 2**). In Mombasa unprocessed/minimally processed were advertised more (52%) than UPF, while in Baringo 64% of the advertisements were UPF.

**Figure 2:**
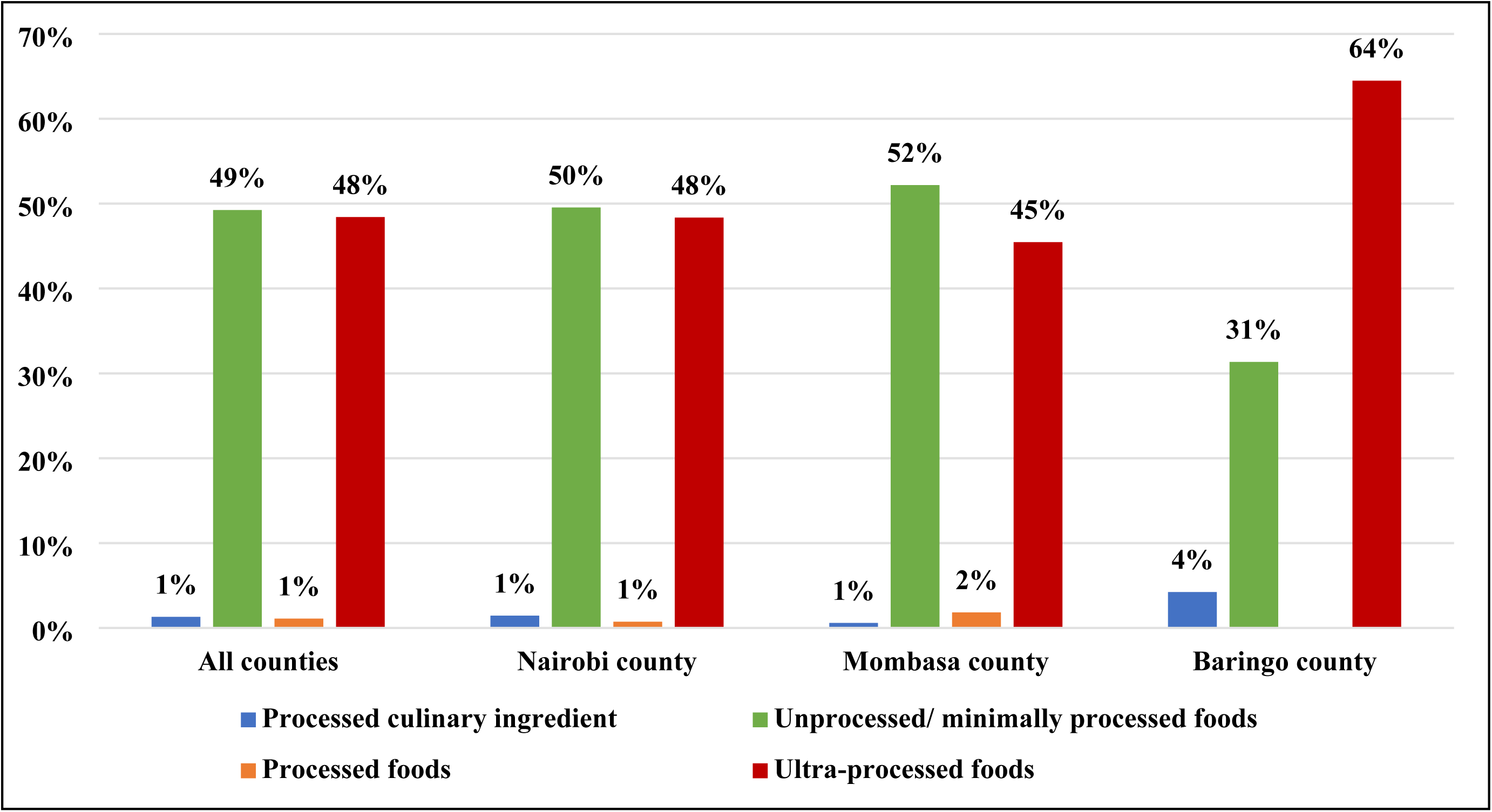
The proportion{%) of advertisedfoods and beverages by NOVAfood categories and counties.

### Description of promotional/marketing strategies in the advertisements

About 62.3% of the advertisements had no promotional character’s present. Among those with promotional characters, cartoon or company-owned characters were the most frequently observed promotional strategy (37.2%). Other observed promotional methods accounting for 2.0% of the advertisements included famous sports persons (n=2), sports events (n=4), movie titles (n=2), and advertisements for children, e.g. images of a child (n=14). The most common premium offers in the advertisements were price discounts (n=49) and limited editions, e.g. last stock (n=33). Other premium offers observed include: contests (n=6) buy one get one free (n=5), gift or collectibles (n=5), and 20% extra (n=4).

### Factors associated with UPF advertisements around schools

There was a significantly higher level of advertisements of UPFs in Baringo County (prevalence rate ratios (PRR): 1.28, 95% CI: 1.01-1.20) compared to urban Nairobi County (**Table 6**). We observed a significantly higher level UPFS advertisements in Lower SES settings (PRR: 1.10, 95% CI: 1.01-1.20).

**Table 6:** Multivariate Poisson regression output for factors associated with levels of advertisements of ultra-processed foods and beverages in Nairobi, Mombasa, and Baringo counties.

|  | *PRR* | *95% CI* | *P-value* |
| --- | --- | --- | --- |
| **County** |  |  |  |
| Ref: Nairobi county | 1 | 1 | 1 |
| Baringo county | 1.28 | [1.00-1.63] | 0.048 |
| Mombasa county | 0.99 | [0.90-1.09] | 0.872 |
| **SES level** |  |  |  |
| Ref: High SES | 1 | 1 | 1 |
| Lower SES | 1.10 | [1.01-1.20] | 0.035 |
| **School Setting** |  |  |  |
| Ref: Urban | 1 | 1 | 1 |
| Rural | 1.05 | [0.83-1.34] | 0.676 |
| **School type** |  |  |  |
| Ref: Primary school | 1 | 1 | 1 |
| Secondary schools | 1.04 | [0.94-1.15] | 0.451 |

Notes: *PRR- Prevalence rate ratios CI-Confidence Interval; SES-Socioeconomic status*

## Discussion

We observed a high level of marketing of UPFs and other unhealthy food and beverages around schools in Kenya, with nearly half of all the food advertisements rated as unhealthy. There was a substantial strategic placement of advertisements around the 250m radius around schools, with the highest numbers being observed in the urban counties (Nairobi and Mombasa) and around primary schools. This is consistent with findings from studies in other countries where there were varying densities of advertisements by SES locations and in urban areas compared to peri-urban neighborhoods (14,18,21). Our findings identified a significantly high level of advertisements in lower-SES areas in the various counties, which is similar to a study conducted in Ghana using the same methodology (18). These findings show that urban settings and both lower and higher SES locations are targeted by food and beverage industries.

Nearly half of the advertisements around schools were in the UPF/unhealthy food category, showing a substantial exposure of children to unhealthy foods, a similar trend reported elsewhere(12,18,21,29). Different studies from LMICs have consistently shown that most advertisements for unhealthy foods strategically target children (12,13,15,30). These child-directed advertisements of unhealthy foods are a great public health concern as they increase the consumption of unhealthy foods which will consequently increase the risk of nutrition-related NCDs (4,31,32).

The most commonly advertised unhealthy food in this study was sugar-sweetened drinks followed by sweetened bread, cakes, and savory snacks. Most studies conducted around schools have shown a high level of promotion of sugar-sweetened beverages to children (18,21). Studies conducted in urban Uganda and Ghana using the INFORMAS methods showed a high level of promotion of unhealthy foods, with sugar-sweetened beverages being the most advertised product (18,21). The high level of exposure to unhealthy food advertisements creates significant risks of poor dietary choices and consequential health-harming implications for children (4,33,34).

Surprisingly, 3% of the advertisements were for alcohol, which is restricted to advertising to children in Kenya (35). Alcohol advertisements around schools have also been observed in Uganda and Ghana, and these advertisements are associated with an increased likelihood of consumption of alcohol by children (18,21,36).

The majority of the advertisements in this study were strategically placed in the food shops/kiosks around the schools and were mostly in the form of posters and banners. Most children purchase both food and non-food products from shops around school, implying that they are exposed to advertisements for unhealthy foods. These findings are consistent with the study conducted in Ghana, where the majority of advertisements around schools were mainly placed in food shops(18). Modern retail outlets and shops are associated with increased exposure and prominence of unhealthy food categories and make different products and brands easily accessible for purchase by children (18,37,38).

Posters and banners are a marketing strategy used to make clear messaging about products, that can be multiply viewed by the target market. As observed in previous studies, physical advertisements using posters and banners improve the proximity and awareness of the food items being advertised and influence the purchase and consumption of these products (18,39,40). This study shows the diversified marketing landscape and its strategic target to vulnerable populations such as children stressing the need for regulations encompassing the marketing strategies, placement, content, and type of advertisements exposed to different populations. The most common promotional strategies used in this study were the cartoon or company-owned characters hence improving the appeal of the advertisements to children. Some studies have shown that the use of cartoon and company-owned characters and mascots enhanced the purchase, and consumption of unhealthy foods, and consequential poor health outcomes (9,10).

### Policy implications

This study illustrates the marketing of UPFs around schools in different geographical contexts in Kenya. The promotion and marketing of unhealthy foods can influence the perception of the product and thus increase the purchase intention of children or pester power to caregivers by children. Consequently, this leads to the consumption of unhealthy foods associated with poor health outcomes such as overweight/obesity, and NR-NCDs among children and adults. The marketing of unhealthy foods, specifically targeting children has been widespread around school neighborhoods. The marketing strategies have been improved and mirror the methods used in HICs as the industrialization and modernization of food environments take their roots in LMICs.

This study contributes to the knowledge of the landscape assessment of marketing around school neighborhoods, with a comparison between rural and urban areas of Kenya. The most worrying observation is the targeting of younger children in primary schools, rural schools, and schools in low SES urban areas. This study calls for a multisectoral approach toward the regulation of unhealthy food advertising in school settings regardless of their location and socio-economic status. There is a need to sensitize school authorities about the dangers of advertising unhealthy foods to children to stimulate local restrictions on such advertisements by the schools. At the school level continuous advocacy, sensitization, and education on predatorial marketing of unhealthy foods, and the health-harming implications should be done to counter the marketing campaigns in school neighborhoods. There needs to be a great focus by the policymakers on the high level of advertisements of sugar-sweetened beverages and alcoholic beverages around schools, as elucidated in this study. The rights-based approach can be implemented as recommended by WHO where it is recommended that marketing around settings where children gather should be free of marketing of unhealthy foods (41).

### Strengths and Limitations

This is the first study in Kenya assessing marketing in neighborhoods where children gather, hence making it novel. This study is unique in that it entails findings of marketing around school neighborhoods from both rural and urban regions in an LMIC setting. Most marketing assessment studies have been conducted in HICs and urban areas of a few countries in Africa. Hence, the study shows a comparative assessment of the Kenyan urban and rural contexts showing a high penetration of UPFs advertisements targeting children. The main limitation of the study was that it was conducted during the COVID-19 period, with numerous restrictions/lockdowns which might have impacted the number of advertisements during the study period. However, most of the advertisements displayed before COVID-19 were available during the survey. So, it is possible that we did not miss a substantial number of advertisements during the study period.

## Conclusion

The findings from this study illustrate the high level of exposure to unhealthy foods and beverages targeting children in both urban and rural settings in Kenya. There is a concerning observation on the use of innovative techniques of marketing around schools, and the widespread advertisement of sugar-sweetened beverages, especially in rural settings. There is an urgent need for the formulation of national policies on the regulation of marketing of unhealthy foods to children, around school environments. Targeted awareness, and sensitization are needed at the school level to inform the children and teachers on the existing marketing of unhealthy foods targeting children.

## Data Availability

The datasets for this study can be available upon reasonable requests by researchers. The data request will undergo data sharing agreement policy review and guidelies at the African Population and Health Research Centre.

## Acknowledgments

The authors acknowledge the Ministries of Health and Education in Kenya, and the county governments of Nairobi, Mombasa, and Baringo for supporting the research in the country and in specific counties. We also acknowledge the research assistants who supported data collection.

